# Maternal DNA Methylation Signatures of Gestational Diabetes across all Stages of Pregnancy

**DOI:** 10.1101/2025.05.25.25328297

**Authors:** Luma Srour, Yosra Bejaoui, Jayakumar Jerobin, Manar Dweik, Aswathy Sankar, Abeer Qannan, Nassima Allouche Colak, Odette Chagoury, Noha A. Yousri, Thomas Farrell, Eleni Fthenou, Mohammed Bashir, Nady El Hajj

**Affiliations:** College of Health and Life Sciences, Hamad Bin Khalifa University, Qatar Foundation, Doha, Qatar; Weill Cornell Medicine - Qatar, Education City, Doha, Qatar; Qatar Metabolic Institute, Hamad Medical Corporation, Doha, Qatar; Department of Research, Women’s Wellness and Research Center, Hamad Medical Corporation, Doha, Qatar; Endocrine Section, Department of Medicine, Hamad Medical Corporation, Doha, Qatar; Computer and Systems Engineering, Alexandria University, Alexandria, Egypt; Qatar Precision Health Institute, Qatar Foundation for Education, Science, and Community, Doha, Qatar; College of Medicine, Qatar University, Doha, Qatar; College of Science and Engineering, Hamad Bin Khalifa University, Qatar Foundation, Doha, Qatar

**Keywords:** Gestational diabetes mellitus, Type 2 diabetes, Cardiovascular disorders, DNA methylation, Qatar Biobank, Qatar Birth Cohort

## Abstract

Gestational diabetes mellitus (GDM) is a common metabolic disorder characterized by hyperglycemia that is first detected during pregnancy, which is not overt diabetes. GDM poses a substantial risk for prenatal and postnatal adverse outcomes affecting both the mother and the offspring. These complications include, but are not limited to, fetal macrosomia, shoulder dystocia, respiratory distress, neonatal hypoglycemia, type 2 diabetes, and cardiovascular diseases. Screening for GDM typically occurs between 24 and 28 weeks of gestation, a timing that is considered late and may increase the risk of all the adverse outcomes associated with GDM. Treatment and prevention strategies are not standardized globally, may be suboptimal, and are often initiated after a diagnosis has been made. Therefore, our primary goal was to identify DNA methylation signatures specific to GDM to understand its underlying mechanisms. We conducted genome-wide DNA methylation profiling for normal and GDM pregnant women across the three trimesters of pregnancy in the discovery cohort. In addition, we validated our findings in a second cohort collected in Qatar. In this study, we uncovered and validated new DNA methylation signatures that may significantly influence the expression of genes associated with GDM. Furthermore, we discovered new genes (*RSL1D1*, *HOXD4*, and *MROH6*) that may play a role in GDM and might be related to the risk of developing T2D and cardiovascular disease later in life. We conclude that DNA methylation changes during pregnancy might not fully explain GDM pathogenesis but can reflect population-specific environmental and behavioral factors before and during pregnancy. Some of these discovered CpG sites might regulate previously reported genes linked to GDM and diabetes, highlighting shared and distinct epigenetic mechanisms across populations.

## Introduction

Gestational diabetes mellitus (GDM) is a metabolic disorder characterized by hyperglycemia. It is primarily detected during pregnancy and usually resolved after delivery [1, 2]. GDM is one of the most common complications associated with pregnancy, affecting approximately 14% of all pregnancies worldwide [3, 4]. Furthermore, GDM poses a substantial risk for prenatal and postnatal adverse outcomes affecting both the mother and the offspring. For the offspring, these risks include conditions such as fetal macrosomia (excessive birth weight), shoulder dystocia, respiratory distress, and neonatal hypoglycemia. For the mother, potential complications include pre-eclampsia, gestational hypertension, type 2 diabetes (T2D), and cardiovascular disease [5]. Screening for diabetes in pregnancy is typically performed between 24-28 weeks of gestation via a standard 75-gram oral glucose tolerance test (OGTT) based on the World Health Organization (WHO) [6]. However, it is arguable that screening at 24-28 weeks of gestation might not be optimal, and earlier screening is required [7]. Early interventions, such as nutritional changes, physical activity, or pharmacological treatment, could significantly improve maternal and child health outcomes [8, 9]. Pharmacological therapies, such as metformin and insulin, are commonly used to manage blood sugar levels in patients. However, oral medications such as metformin can cross the placenta and may impact fetal development. Although insulin is a safer option, it has low effectiveness due to the current state of insulin resistance in GDM patients and involves higher costs and necessitates patient education [10, 11]. Therefore, there is an urgent need for safe and effective treatment of GDM, as well as the development of tests for the early detection of GDM. Achieving this requires a better understanding of GDM pathogenesis and underlying mechanisms.

Although the pathogenesis of GDM is not yet fully understood, it is recognized as a multifactorial disorder influenced by genetic and epigenetic factors, environmental exposures, inflammatory factors, and oxidative stress [11–13]. It is known that a complex interplay of risk factors influences GDM development; for example, obesity and physical inactivity are prominent factors, with a sedentary lifestyle ranking as one of the leading contributors affecting women’s overall health [14–16]. In addition, advanced maternal age, multiparity, a family history of T2D, and specific obstetric and medical factors, such as a history of delivering a child with macrosomia, previous GDM history, and polycystic ovary syndrome, further elevate the risk of GDM [17–22]. During early pregnancy, the body starts to prepare for the vast energy demand by increasing glucose uptake and storage in adipose tissues by significantly increasing insulin sensitivity. As pregnancy progresses, various local and placental hormones, such as lactogen, estrogen, and prolactin, rise in a coordinated manner, leading to increased insulin resistance. This shift is advantageous as it helps maintain higher glucose levels in the blood, ensuring enough glucose is available for the fetus by allowing it to be transported through the placenta [11]. GDM usually develops due to a defect in insulin secretion, an insulin sensitivity defect, or both [23]. GDM is a heterogeneous condition with different subgroups, severity levels, and adverse outcomes influenced by genetic/epigenetic, environmental factors, metabolic profiles, and comorbidities [11, 23–25]. Studying each factor contributing to GDM is essential for understanding this diversity, which can help improve risk assessment, prevention, treatment strategies, and the identification of early biomarkers for GDM prediction.

Epigenetic mechanisms, including histone modifications and DNA methylation, are known to be implicated in GDM pathogenesis. Epigenetics is considered the bridge between genetics and the environment, as it can explain the effect of environmental factors on disease development [26]. Recently, different studies have explored the role of epigenetic modifications in GDM, with DNA methylation emerging as one of the most studied mechanisms. However, most studies have focused on offspring exposed to a diabetic *in utero* environment [13, 27, 28]. Few studies have explored the effects of GDM on maternal DNA methylation signatures of pregnant women, mainly in European and South Asian populations [29, 30]. It is essential to note that methylation signatures can vary between populations due to differences in behavior and environmental factors [31]. Epigenome-wide association studies (EWAS) on GDM have not been previously conducted in Qatar, despite GDM being a significant global health issue that affects approximately 31.6% of all pregnancies [3, 4, 32]. This is due to the high prevalence of obesity (70.1%) and diabetes (16.7%) in the country.

In this study, we conducted a genome-wide DNA methylation screening in pregnant women recruited as part of the Qatar Birth Cohort (QBiC) study. Then we validated our findings in a second cohort recruited at the Women’s Wellness and Research Center (WWRC). We aimed to elucidate the role of DNA methylation in the development of GDM and understand the pathways and epigenetic changes associated with this condition across the three trimesters of pregnancy in longitudinally followed pregnancies. Furthermore, we aimed to uncover genes and behavioral and environmental factors that might drive GDM pathogenesis. Moreover, we compared GDM-related DNA methylation changes to those observed in females with Type 2 Diabetes (T2D) to identify epigenetic alterations that may predispose women affected by GDM to develop T2D later in life.

## Materials and methods

### Discovery cohort

Our discovery cohort included pregnant women (N=140), a subset of the Qatar Birth Cohort (QBiC) study, recruited at Qatar Biobank (QBB) between April 2019 and December 2023 [33]. QBiC cohort enrolled adult (age≥ 18 years) pregnant women, Qatari or women who lived in Qatar for at least 15 years, who live in Qatar and plan to give birth in Qatar, who speak or understand English or Arabic. For this study, 66 GDM pregnant women and 74 non-GDM pregnant women (controls) were involved. Women with pre-existing diabetes history and controls with polycystic ovary syndrome (PCOS) were excluded from the differential methylation analysis. A sample of 61 GDM women and 63 controls were examined for differential DNA methylation analysis.

Informed consent was obtained from all the participants. The GDM women were identified by self-reported questionnaires to be diagnosed by a doctor. Height and pre-pregnancy weight were measured at T0 to calculate ‘pre-pregnant body mass index (BMI)’ as weight (kg)/ height^2^ (m^2^). Weight gain was calculated as the difference between the pre-pregnancy weight at T0 and baseline visit T1 (1-13 weeks), T0 and T2, and T0 and T3.

All participants’ information was collected via electronic questionnaires related to diet during pregnancy, lifestyle, environmental exposure, medical history, mental health, pregnancy history, family history, and previous pregnancies. Biological samples were collected, such as whole blood, saliva, urine, and stool samples. Biochemical, Hematology, endocrinology, and clinical chemistry laboratory test results were performed at Hamad Medical Corporation (the main public Healthcare provider in Qatar).

This study was approved by the institutional review boards (IRB) of QBB (Ex-2022-QF-QBB-RES-ACC-00100-0203) and Hamad Bin Khalifa University with approval number: QBRI-IRB 2021-09-107.

### Validation Cohort

The validation cohort was recruited from the Endocrine and Pregnancy Unit at the Women’s Wellness and Research Centre (WWRC) at Hamad Medical Corporation (HMC), which is the main national hospital and provider of secondary and tertiary health care facilities in Qatar, in the period between December 2022 and April 2024. Qatar also implements a national screening program for diabetes in pregnancy [32]. The inclusion criteria were pregnant women aged 18–50 years and being in the 24-28 weeks of pregnancy. Thirty-five pregnant women with GDM and 37 non-GDM pregnant women (controls) were studied. The GDM diagnosis was made as follows: firstly, all high-risk women without pre-existing diabetes mellitus (DM) are screened using fasting plasma glucose and HbA1c tests. If the initial test results are normal, high-risk women undergo a 75g OGTT at 16–18 weeks of gestation. The OGTT is repeated at 24–28 weeks of gestation if the earlier test is also normal. Low-risk women are screened at 24–28 weeks with the same 75g OGTT. For OGTT, GDM is defined based on the WHO-2013 criteria: FBG 5.1–6.9 mmoL/l, 1 hour ≥10.0 mmoL/l, and 2 hours 8.5–11.0 mmol/L. Those pregnant women with normal OGTT were considered as controls (non-GDM).

The study protocol includes a baseline visit (at the time of performance of OGTT, at 24– 28 weeks of pregnancy; T2), a pre-labor visit (at 34–38 weeks of pregnancy; T3). Informed consent was obtained from all the participants in this study in accordance with the Declaration of Helsinki. The Medical Research Center (MRC), Hamad Medical Corporation, approved the study with the approval number: MRC-03-22-119.

At the Endocrine and Pregnancy Unit, height was measured at T0, and weight and blood pressure (BP) were measured at every visit. At T0, pre-pregnancy weight was collected to calculate the pre-pregnancy BMI. Weight gain was calculated as the difference between the pre-pregnancy weight at T0 and baseline visit T2 (24–28 weeks) and between T0 and T3. All participants’ information was collected via a standardized questionnaire that included general and specialized questions, medical history, pregnancy history, family history of diabetes, and previous pregnancies. Several medical and clinical variables were obtained from the electronic health records (an integrated electronic health record including all inpatient and outpatient visits at the WWRC). Clinical data, biochemical variables, laboratory test results, and medical history were collected using the electronic medical records at WWRC.

Women were excluded from this study if they were unable to provide informed consent, were newly diagnosed with T2D, on Metformin therapy or steroid treatment, had known or suspected fetal congenital malformations or chromosomal abnormalities, inability or refusal to complete the oral glucose tolerance test (OGTT) due to nausea or vomiting, planning to deliver at a different healthcare facility, participation in another clinical trial, a history of bariatric surgery, or failure to complete all required study procedures.

### DNA Methylation Profiling

According to the manufacturer’s instructions, DNA methylation was profiled via the Infinium MethylationEPIC v2.0 BeadChip. From each sample, 500 ng was used for bisulfite conversion via the EZ DNA Methylation Kit (Zymo Research, Irvine, CA, USA), followed by DNA amplification, fragmentation, and hybridization to the BeadChip. The extension process is then performed with labeled nucleotides, and finally, the arrays are scanned via the Illumina iScan to measure methylation levels. The raw intensity data files (IDATs) were analyzed via the RnBeads package in R Studio (version 4.4.2). The analysis pipeline included a data quality check, preprocessing, blood cell estimation, and differential DNA methylation analysis. The preprocessing steps included filtering out probes that overlapped with SNPs and cross-reactive probes and removing samples and probes with the highest fraction of unreliable measurements via the GreedyCut algorithm, in addition to normalization via the beta-mixture quantile normalization method (BMIQ). Probes found on sex chromosomes were additionally removed during preprocessing. The blood cell proportion was estimated using the FlowSorted.Blood.EPIC package. Finally, differential methylation analysis was conducted separately for each trimester at both the CpG site and region levels (regions included promoters, CpG islands, and tiling in 5 kb windows). Differential methylation analysis is based on a linear model from the limma package. In the linear model, we adjusted for the following covariates: age, BMI, treatment, blood cell proportions, and batch effect. For differential methylation analysis, M-values (logit-transformed beta values) were used. The comparisons in the differential methylation of the discovery cohort were as follows: 15 cases vs. 11 controls in T1, 36 cases vs. 42 controls in T2, and 59 cases vs. 58 controls in T3. The comparisons in the validation cohort were 33 cases vs. 37 controls in T2 and 33 cases vs. 37 controls in T3.

### Validation of Differentially Methylated Sites and Regions

Using Venny 2.0 software, common differentially methylated probes/sites (DMPs) and regions (DMRs) were identified between the discovery and validation cohorts. To validate the DMPs found in both the T2 and T3 cohorts, we used the top 10,000 combined-rank CpG sites from the QBiC and WWRC cohorts. We additionally examined the overlap between the top 10,000 combined-rank DMPs at T1 from the QBiC and the validated DMPs identified at both T2 and T3. Furthermore, the top 100 combined-rank regions, including promoters, CpG islands, and tiling, were utilized to validate our findings at the regional level. For further analysis, we focused on DMPs and differentially methylated regions (DMRs) that were either hypermethylated or hypomethylated in both cohorts. CpG sites associated with validated promoters were obtained from Illumina’s Infinium MethylationEPIC v2.0 Manifest. Finally, the GREAT tool (version 4.0.4) was used to annotate the noncoding genomic regions (CpG islands and tiling) to their nearest genes.

### Identification of Differentially Methylated Sites Associated with T2D

Linear regression analysis was conducted to assess the association between age-corrected CpG methylation levels and T2D status in 609 females recruited by QBB as part of a study conducted by Yousri *et al*. in 2023 [34]. The model included the following covariates: BMI, the first two principal components (PC1 and PC2) derived from measured cell counts (neutrophils, basophils, eosinophils, monocytes, lymphocytes), sample plate, well position, a smoking surrogate based on AHRR methylation, three genomic principal components from whole-genome data, and batch effect. Gene annotation of the identified CpG sites was guided by the Illumina MethylationEPIC v2.0 array manifest.

### Downstream Analysis

Downstream analysis was conducted via the methylGSA package for KEGG pathway and Gene Ontology (GO) enrichment analysis, and eFORGE 2.0 for tissue- or cell type-specific signal identification. Downstream analysis was performed for validated CpG sites in T2 and T3 only. A specific annotation package (IlluminaHumanMethylationEPICv2anno.20a1.hg38) for EPIC v2 was used during the KEGG pathway and GO enrichment analyses. eFORGE 2.0 software was used on validated DMPs across different trimesters to identify tissue-specific signals by overlapping these sites with DNase I hypersensitive site (DHS) data from ENCODE. In addition, the association of the validated DMPs with known biological traits was analyzed via the EWAS toolkit. The effect of methylation changes in the promoter region on gene expression was assessed using the EWAS toolkit (https://ngdc.cncb.ac.cn/ewas/toolkit, accessed 27 August 2024) [35]. To further validate the effects of DNA methylation alterations on the gene expression of nearby genes, methylation quantitative trait loci (meQTLs) were examined via the BIOS QTL browser [36]. In addition, traits linked to the identified SNPs were checked using the GWAS catalog [37],

### Transcriptomic validation of epigenetically dysregulated candidate genes in GDM

To study the effect of changes in DNA methylation on gene expression, we used a publicly available transcriptomic dataset (GSE255075) generated using high-throughput sequencing by Illumina NovaSeq 6000. This dataset was obtained from placental tissue samples collected from normal and GDM singleton-term pregnant women during cesarean deliveries at the Affiliated Hospital of Jining Medical University in China. Raw fragment counts for normal (n =3) and GDM (n =3) samples were downloaded from the Gene Expression Omnibus (GEO) database. Differential gene expression analysis was conducted in R Studio (version 4.4.2) using the edgeR package.

### Statistical testing

All the statistical tests were performed using R Studio (version 4.4.2). Continuous variables are presented as mean ± standard deviation or median and inter-quartile range, where appropriate, whereas categorical data are presented as a percentage. Student t-test and Mann-Whitney U test were used to compare continuous variables, while categorical variables were compared using the Chi-square test. For data that followed a normal distribution, the student’s t-test was used to compute the significant differences in body composition metrics in the QBiC cohort, while it was used for several metabolic parameters measured in the WWRC cohort, including maternal age, fasting blood glucose, OGTT results (after 1 hour and 2 hours), and HbA1c%. Conversely, the nonparametric Mann‒Whitney U test was employed for data that did not meet the normal distribution assumption. In the discovery cohort, the Mann‒Whitney U test was used for maternal age, insulin levels, and HbA1c. In the validation cohort, all body composition metrics followed a skewed distribution. The chi-square test was used to assess significant differences in nationality between the groups in both cohorts. In addition to significant differences in juice consumption, coffee consumption, smoking, smoking exposure, and exercise activity.

## Results

### Anthropometric and metabolic parameters of the studied cohorts

To evaluate the differences between the two groups (cases and controls) in the discovery cohort, we analyzed various anthropometric and metabolic parameters, including maternal age, weight, BMI, insulin, and HbA1c levels. Our analysis revealed substantial differences between GDM cases and controls, as shown in Table 1. The GDM group had a significantly greater pre-pregnancy BMI (29.8 ± 5.1 kg/m², p-value = 0.0011) than the control group (26.8 ± 5.1 kg/m²). Similarly, GDM patients had a higher BMI during pregnancy at T1 (30.2 ± 4.5 kg/m², p-value = 0.037), T2 (31 ± 5.6 kg/m², p-value = 0.023), and T3 (32.6 ± 5 kg/m², p-value = 0.027) compared to the controls (T1: 26.8 ± 5.5 kg/m², T2: 28.5 ± 5.1 kg/m², T3: 30.7 ± 5 kg/m²). Insulin levels also displayed significant differences at T1 (GDM median of 15.8 mcunit/mL and control median of 7.3, p-value = 0.022) and T2 (12.2 mcunit/mL in GDM patients and 8.2 mcunit/mL in controls, p-value = 0.017); however, at T3, the levels (18.4 mcunit/mL) were similar to those of controls (14.6 mcunit/mL, p-value = 0.28). Similarly, the difference in HbA1c levels tended to increase from T1 (p-value = 0.0033) to T2, with a more pronounced difference at T2 (p-value = 8.12e-05), followed by a decline at T3 (p-value = 0.081). Maternal age and nationalities were similar in both groups.

**Table 1:** Anthropometric and metabolic characteristics of the QBiC GDM cases and controls.

| QBiC (Discovery Cohort) |  |  |  |  |
| --- | --- | --- | --- | --- |
|  |  | GDM (n = 66) | Control (n = 74) | p-value |
| Maternal Age |  | (30.5, 27 – 35.8) (n = 66) | (29, 26 - 36) (n = 74) | 0.33 |
| Pre-pregnancy BMI (kg/m<sup>2</sup>) | | $29.8 \pm 5.1$ (n = 60) | $26.8 \pm 5.1$ (n = 74) | 0.0011 |
| Pre-pregnancy weight (kg) | | $75.8 \pm 14.4$ (n = 61) | $68.1 \pm 12.8$ (n = 74) | 0.0012 |
| Gestational weight gain (kg) | T1 | $0.8 \pm 4.1$ (n = 19) | $1.8 \pm 4.1$ (n = 19) | 0.48 |
| | T2 | $1.8 \pm 6$ (n = 41) | $3.9 \pm 5$ (n = 55) | 0.065 |
| | T3 | $6.2 \pm 7.2$ (n = 60) | $10 \pm 6.3$ (n = 74) | 0.0016 |
| Insulin (mcunit/mL) | T1 | (15.8, 7.6 - 39.8) (n = 21) | (7.3, 5.4 - 13.8) (n = 19) | 0.022 |
|  | T2 | (12.2, 9 - 23.7) (n = 44) | (8.2, 5.9 - 14) (n = 54) | 0.017 |
|  | T3 | (18.4, 10.9 - 32.8) (n = 63) | (14.6, 9.4 - 31.3) (n = 70) | 0.28 |
| HOMA-IR | T1 | (2.8, 1.6 - 9.7) (n = 21) | (1.3, 1.1 - 2.3) (n = 19) | 0.017 |
|  | T2 | (2.6, 1.8 - 5) (n = 44) | (1.6, 1.1 - 2.7) (n = 54) | 0.0078 |
|  | T3 | (5.3, 5.2 - 5.5) (n = 21) | (5, 4.9 - 5.2) (n = 19) | 0.16 |
| HbA1C % | T1 | (5.3, 5.2 - 5.5) (n = 21) | (5, 4.9 - 5.2) (n = 19) | 0.0033 |
|  | T2 | (5.3, 5.1 - 5.6) (n = 44) | (5, 4.8 - 5.2) (n = 54) | < 0.001 |
|  | T3 | (5.3, 5.1 – 5.5) (n = 63) | (5.2, 5 – 5.4) (n = 70) | 0.081 |

Anthropometric and metabolic parameters, including maternal age, nationality, and body composition metrics, were comparable between the GDM and non-GDM groups in the validation cohort. However, metabolic parameters, including fasting blood glucose, OGTT, and HbA1C, were significantly different, as indicated in Table 2. Fasting blood glucose was significantly elevated in the GDM group (5 ± 0.4 mmol/L, p-value = 3.33e-08) compared with the control group (4.5 ± 0.3 mmol/L). Evidently, OGTTs after 1 hour and 2 hours levels were higher in GDM patients (1 hour: 10.3 ± 1.2, 2 hours: 8.2 ± 1.6) than in controls (1 hour: 7.4 ± 1.2, 2 hours: 6.2 ± 1.4) with p-value = 8.94e-15 and p-value = 5.88e-07 after 1 hour and 2 hours, respectively. Finally, GDM HbA1c levels were significantly higher (5.4 ± 0.4, p-value = 0.0039) than those of controls (5.1 ± 0.4).

**Table 2:**
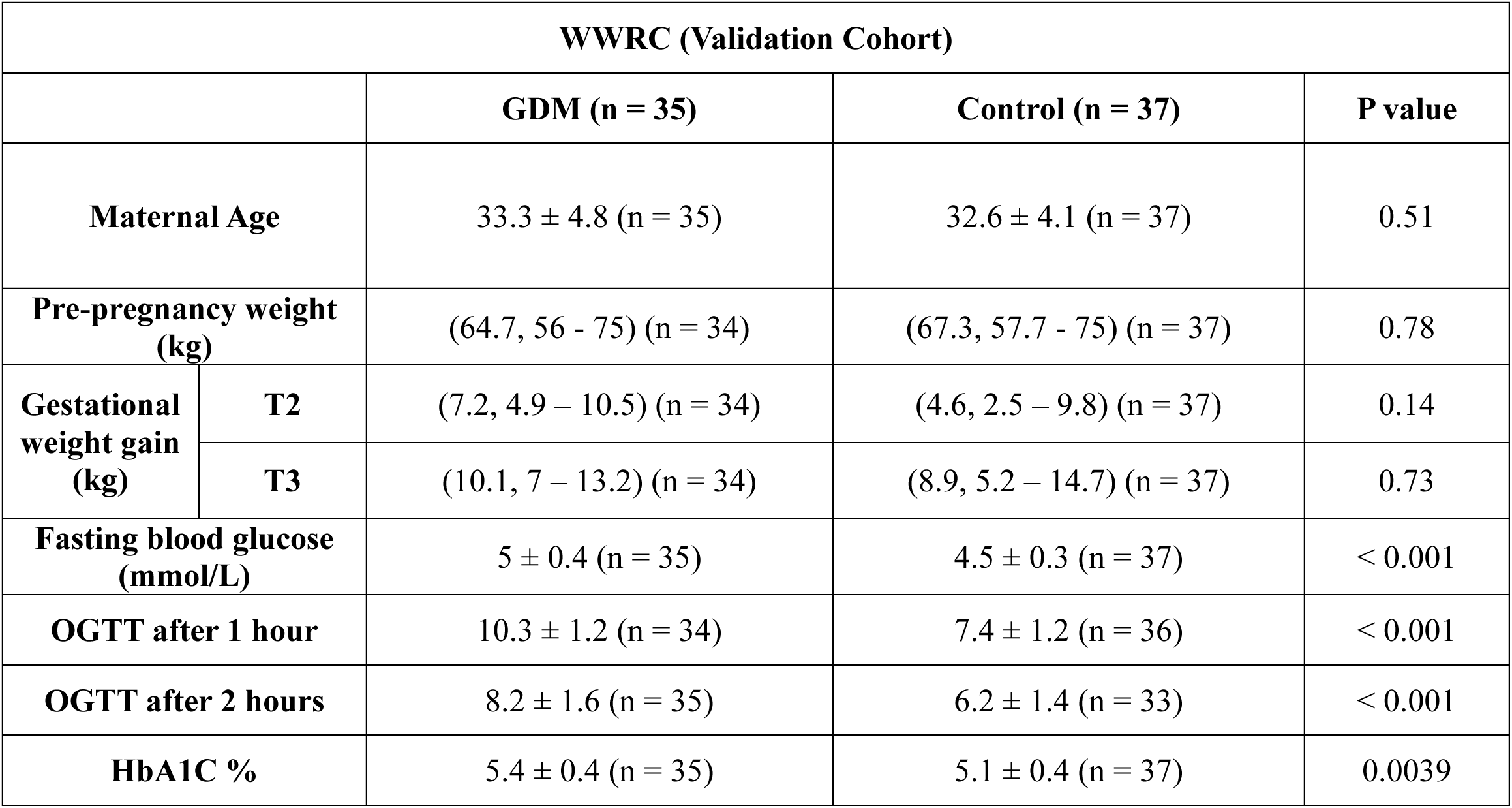
Anthropometric and metabolic characteristics of the Women’s Wellness and Research Centre cohort.

| WWRC (Validation Cohort) |  |  |  |  |
| --- | --- | --- | --- | --- |
|  |  | GDM (n = 35) | Control (n = 37) | P value |
| Maternal Age |  | 33.3 ± 4.8 (n = 35) | 32.6 ± 4.1 (n = 37) | 0.51 |
| Pre-pregnancy weight (kg) |  | (64.7, 56 - 75) (n = 34) | (67.3, 57.7 - 75) (n = 37) | 0.78 |
| Gestational weight gain (kg) | T2 | (7.2, 4.9 – 10.5) (n = 34) | (4.6, 2.5 – 9.8) (n = 37) | 0.14 |
|  | T3 | (10.1, 7 – 13.2) (n = 34) | (8.9, 5.2 – 14.7) (n = 37) | 0.73 |
| Fasting blood glucose (mmol/L) |  | 5 ± 0.4 (n = 35) | 4.5 ± 0.3 (n = 37) | < 0.001 |
| OGTT after 1 hour |  | 10.3 ± 1.2 (n = 34) | 7.4 ± 1.2 (n = 36) | < 0.001 |
| OGTT after 2 hours |  | 8.2 ± 1.6 (n = 35) | 6.2 ± 1.4 (n = 33) | < 0.001 |
| HbA1C % |  | 5.4 ± 0.4 (n = 35) | 5.1 ± 0.4 (n = 37) | 0.0039 |

### Differentially Methylated Sites across Different Trimesters of Pregnancy

Differentially methylated sites were identified for the GDM group vs controls in each trimester for the discovery and validation cohorts via the RnBeads package. During the preprocessing of the raw data from WWRC, 9981 probes overlapping SNPs were removed, and 30339 probes and 1 sample were filtered out via the GreedyCut algorithm. The remaining samples included 37 controls at two time points (T2 and T3) and 33 GDM women at two time points (T2 and T3), for a total of 140 samples. On the other hand, 9981 probes overlapping SNPs were removed, and 20744 probes were filtered out via the GreedyCut algorithm from the raw QBiC data. The top 10,000 combined-rank CpG sites from T2 and T3 in the QBiC and WWRC cohorts were obtained to validate our findings of the discovery cohort. The combined rank is calculated based on the mean methylation level difference between cases and controls, the quotient of the mean methylation, and the p-value of the statistical test conducted by limma. A smaller (lower) rank indicates stronger evidence of differential methylation between cases and controls. In the WWRC cohort, 404 differentially methylated probes (DMPs) were validated at time point T2, while 536 DMPs were validated at time point T3, as illustrated in Figure 1A. Out of the 404 CpG sites identified at T2, 329 were specific to that time point. Similarly, from the 536 DMPs at T3, 428 were specific to T3, as shown in Supplementary Figure 1. Among the 329 sites in T2, 110 were hypermethylated, and 51 were hypomethylated in both cohorts. Moreover, in T3, 16 sites were hypermethylated, and 184 sites were hypomethylated in the discovery and validation cohorts. Five validated CpG sites in T2 were common with DMPs identified in the first trimester from the QBiC cohort, and all these sites were hypermethylated, as indicated in Table 3. Furthermore, two validated CpG sites in T3 were common with T1 DMPs from the QBiC cohort, out of which one was hypermethylated, and the other was hypomethylated (Table 4).

**Figure 1.**
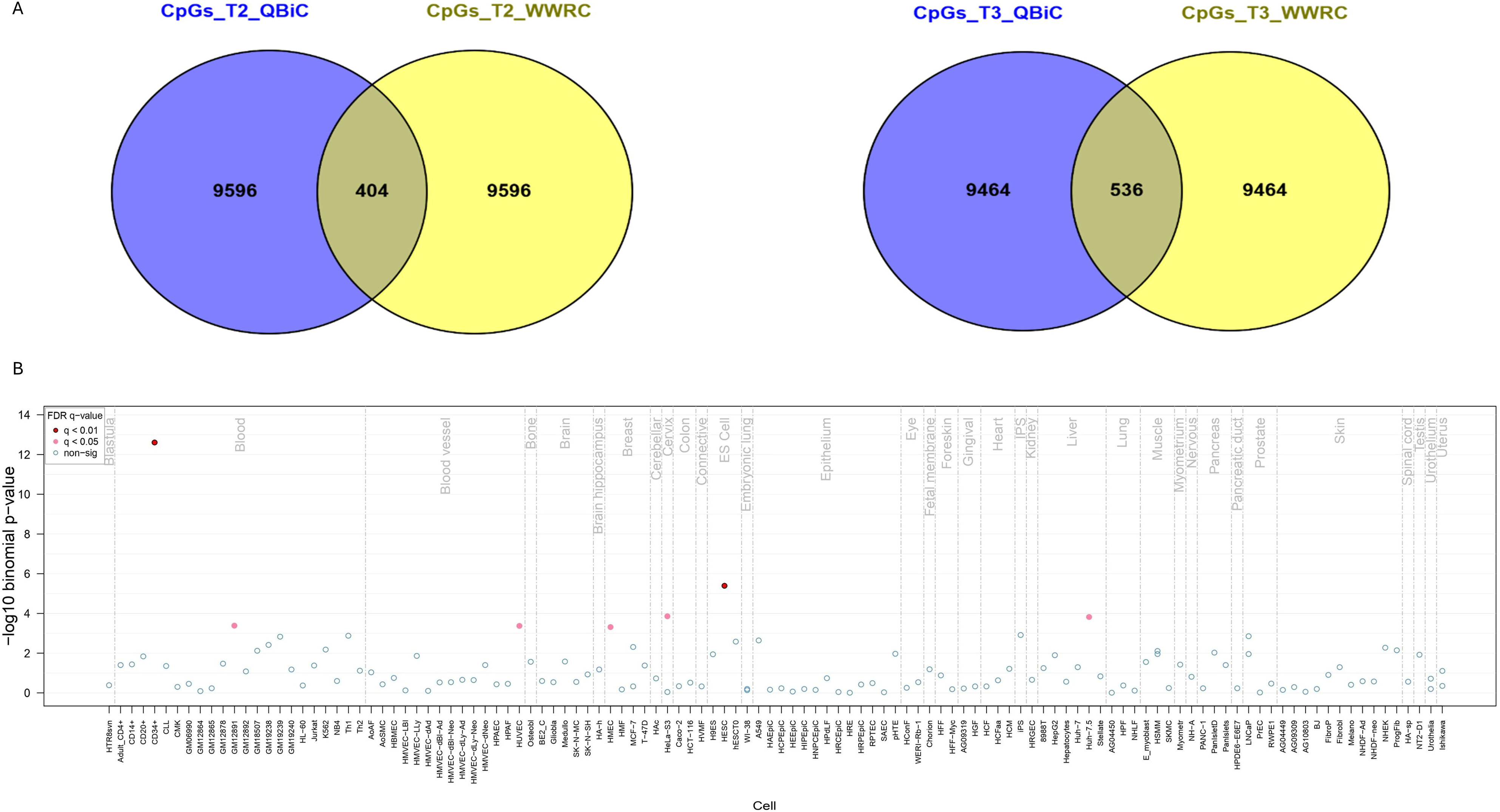
(A) Venn diagram showing the top 10,000 combined ranks in Trimesters 2 and 3 from the QBiC and WWRC cohorts. (B) eFORGE tissue-specific signals of the hypomethylated CpG sites in Trimester 3.

**Table 3:** Differentially methylated CpG sites overlapping between T1 and validated CpGs at T2.

| Cgid | Id | Chromosome | Start | Category |
| --- | --- | --- | --- | --- |
| cg11366711 | 550919 | chr11 | 130313120 | hypermethylated |
| cg15793613 | 95231 | chr2 | 29315462 | hypermethylated |
| cg25306586 | 369572 | chr7 | 103557892 | hypermethylated |
| cg26229990 | 624394 | chr14 | 24332095 | hypermethylated |
| cg27621864 | 138590 | chr2 | 215848049 | hypermethylated |

**Table 4:** Differentially methylated CpG sites overlapping between T1 and validated CpGs at T3.

| Cgid | Id | Chromosome | Start | Category |
| --- | --- | --- | --- | --- |
| cg01075796 | 216398 | chr4 | 54105086 | hypomethylated |
| cg09656629 | 335256 | chr6 | 162151058 | hypermethylated |

### Differentially Methylated Regions across Different Trimesters

Among the top 100 combined-ranked DMRs, one hypermethylated promoter (mean difference > 2%) in T2 was associated with the *AGBL3* gene located on chromosome 7 (chr7:134985008_134987007). According to the Illumina methylation EPIC array v2.0 manifest and RnBeads, this region is associated with 10 CpG sites (cg07277447, cg10731606, cg21033514, cg23733562, cg24097460, cg05351601, cg05813365, cg00588300, cg02015280, and cg03385973). Additionally, we discovered three validated CpG islands in T2 located at chr6:165854488-165854973, chr20:63454211-63454591, and chr17:36703964-36704247.

However, these islands presented a beta methylation difference of < 2% and were associated with only one or two CpG sites each. All these noncoding regions were annotated via GREAT to their nearest genes using the associated genomic regions option. As a result, chr17:36703964-36704247 was annotated to the *LHX1* gene with a distance of 233,369 bases upstream of the transcription start site (TSS). The *LHX1* gene encodes a transcription factor that binds to DNA through a specific domain called the LIM domain, which is a unique zinc-binding domain rich in cysteine. The *LHX1* gene has been implicated in maturity-onset diabetes of the young [38].

### Test Association of Genome-Wide DNA Methylation in GDM Women and T2D Female Patients

Our analysis identified four differentially methylated CpG sites in T2 (cg17207772, cg07998866, cg06392883, and cg00271766) and one site in T3 (cg10113760) associated with GDM and replicated in T2D female patients. These sites have a beta difference of > 2% and a nominal p-value < 0.05. According to the Illumina methylation EPIC array v2.0 manifest, cg07998866 site is located within 200 base pairs upstream of the TSS of the *RGS4* gene. Similarly, the cg06392883 site is located within 1500 bp upstream of the TSS of the *CARD8* gene, while the cg10113760 site is found within 1500 bp upstream of the TSS of the *LINC02135* gene.

### KEGG pathway and GO enrichment analyses

KEGG pathway analysis revealed a few pathways associated with hypermethylated sites with a nominal p-value < 0.05. However, none survived FDR adjustment. As shown in Table 5, these pathways included mannose-type O-glycan biosynthesis and taste transduction. Furthermore, the Rap1 signaling pathway was identified, however this pathway did not reach statistical significance.

**Table 5:** KEGG pathway analysis for validated CpG sites in T2.

| Category | Entry | Description | N | DE | P.DE |
| --- | --- | --- | --- | --- | --- |
| **Hypermethylated CpG sites** | hsa00515 | Mannose type O-glycan biosynthesis | 23 | 1 | 0.008971 |
|  | hsa04742 | Taste transduction | 85 | 1 | 0.031446 |
|  | hsa04015 | Rap1 signaling pathway | 211 | 1 | 0.080382 |
| **Hypomethylated CpG sites** | hsa04144 | Endocytosis | 245 | 1 | 0.072777 |

Gene Ontology (GO) enrichment analysis was similarly conducted in R Studio using the MissMethyl package for the hypermethylated and hypomethylated CpG sites in T2. This analysis revealed 23 and 39 biological pathways with nominal p-values < 0.05 associated with hypermethylated and hypomethylated CpG sites, respectively (Supplementary Tables 1 and 2). The most significantly enriched GO terms related to hypermethylated sites were associated with glycoprotein biosynthetic process, heart growth and development, and sensory perception of taste. Furthermore, pathways related to viral budding via the host ESCRT complex, transport, ribosomal large subunit biogenesis, endomembrane system organization, protein localization, cellular senescence, and smooth muscle contraction were identified from hypomethylated CpG sites.

Furthermore, we employed the MissMethyl package to identify the KEGG pathways and GO terms associated with DMPs in T3. Several pathways were found to be associated with hypomethylated sites at nominal levels. Nevertheless, none survived FDR adjustment (Table 6). These pathways included arrhythmogenic right ventricular cardiomyopathy, hypertrophic cardiomyopathy, dilated cardiomyopathy, taurine and hypotaurine metabolism, butanoate metabolism, beta-alanine metabolism, type I diabetes mellitus, and alanine, aspartate, and glutamate metabolism. However, no KEGG pathways associated with hypermethylated regions were identified. Moreover, the most significant GO terms were identified from hypomethylated sites in T3 (Supplementary Tables 3 and 4). These pathways were related to the regulation of action potential, response to stimulus, metabolic process, T-helper cell differentiation, protein targeting to the mitochondrion, protein-containing complex assembly, protein localization to the mitochondrion, gene expression, glycerolipid biosynthetic process, calcium ion transport, homeostasis, type 2 immune response, and developmental processes.

**Table 6:** KEGG pathway analysis for validated CpG sites in T3.

| **Hypomethylated CpG sites** |  |  |  |  |
| --- | --- | --- | --- | --- |
| **Entry** | **Description** | **N** | **DE** | **P.DE** |
| hsa05412 | Arrhythmogenic right ventricular cardiomyopathy | 86 | 2 | 0.005872 |
| hsa05410 | Hypertrophic cardiomyopathy | 99 | 2 | 0.007345 |
| hsa05414 | Dilated cardiomyopathy | 104 | 2 | 0.008318 |
| hsa00430 | Taurine and hypotaurine metabolism | 16 | 1 | 0.015888 |
| hsa00650 | Butanoate metabolism | 25 | 1 | 0.025752 |
| hsa00410 | beta-Alanine metabolism | 31 | 1 | 0.035935 |
| hsa04940 | Type I diabetes mellitus | 40 | 1 | 0.042556 |
| hsa00250 | Alanine, aspartate and glutamate metabolism | 37 | 1 | 0.042735 |
| hsa04961 | Endocrine and other factor-regulated calcium reabsorption | 53 | 1 | 0.065548 |
| hsa04978 | Mineral absorption | 60 | 1 | 0.067725 |
| hsa05321 | Inflammatory bowel disease | 62 | 1 | 0.069968 |
| hsa00561 | Glycerolipid metabolism | 63 | 1 | 0.071515 |
| hsa04260 | Cardiac muscle contraction | 83 | 1 | 0.094271 |

### Tissue-specific signal identification

The eFORGE tool was used on the list of DMPs from T2 and T3 to identify cell type-specific signals. EPIC v2 probes overlapping with EPIC v1 probes were used as inputs for eFORGE. The tissue-specific signal was identified for the common hypomethylated sites in T3, as shown in Figure 1B. The most significant signal with an FDR < 0.01 was detected in blood and embryonic stem cells (ES cells). eFORGE analysis also revealed significant enrichment (FDR <0.05) of hypomethylated CpG sites in blood, blood vessels, breast, cervix, and liver.

### Trait Enrichment and Expression Regulation

To identify biological traits associated with DMPs, the EWAS toolkit (https://ngdc.cncb.ac.cn/ewas/toolkit, accessed 27 August 2024) was used on the validated DMS (hypermethylated and hypomethylated CpG sites) from T2 and T3. The top biological traits associated with the hypermethylated sites at T2 included atherosclerosis, preterm birth, leukoaraiosis, smoking, vitamin B12 supplementation, glucocorticoid exposure, aging, and exercise, as shown in Supplementary Table 5. On the other hand, the top detected traits associated with the hypomethylated sites investigated at T2 included maternal smoking, leukoaraiosis, in-utero arsenic exposure, and smoking (Supplementary Table 6). Furthermore, traits associated with hypomethylated CpGs at T3 were related to aging, maternal smoking, smoking, juice consumption, blood protein biomarker levels, body mass index (BMI), and other factors (Supplementary Table 7). However, no traits were associated with hypermethylated CpG sites in T3. We analyzed the clinical data collected from the QBiC study to investigate lifestyle-related traits. Out of 62 participants (the total number of responders from the GDM participants), only 15 patients did not consume juice during their pregnancy, while most reported varying frequencies of juice consumption. Out of 65 non-GDM, only 8 did not consume juice during their pregnancy, while most reported varying frequencies of juice consumption. Using the Chi-square test, we found that there is no significant difference (p-value = 0.13) in juice consumption between GDM and non-GDM. Additionally, only a small number of GDM, 8 out of 66 (0.12), smoked at some point in their lives, and only one individual smoked during pregnancy. Of the non-GDM participants, 6 out of 74 smoked at some point in their lives, and none of the controls smoked during pregnancy. There was no significant association between case-control status and lifetime smoking, as determined by the chi-square test (p-value = 0.61). Surprisingly, 35 out of 66 GDM patients were regularly exposed to smoke due to the presence of smokers in their household, while only 16 out of 74 non-GDM were regularly exposed to smoke. The chi-square test results show a significant association between case-control status and household smoke exposure (p-value = 0.00023). We also examined exercise activity, but only 9 out of 66 GDM participants and 15 out of 74 non-GDM engaged in exercise during pregnancy, while 85% of GDM participants (56/66) and 84% of controls (62/74) were involved in other work activities. However, exercise and other working activities were not significantly different between cases and controls.

In addition to trait enrichment analysis, gene expression regulation was explored via the EWAS toolkit. We found that changes in DNA methylation at CpG sites located in the promoter region significantly impact the regulation of gene expression across six tissues (brain, colon, kidney, liver, stomach, and testis), as shown in Figure 2. This analysis revealed several strong negative correlations, suggesting a negative relationship between the methylation level of the input probes and the expression of nearby genes. For instance, in T2, PRSS22_cg21452411 and PRSS22_cg05895034 showed consistent negative correlations in kidney and stomach tissues. Notably, RALGAPA1_cg21911968 exhibited a strong negative correlation in the testis. In contrast, PLEKHG2_cg07774964 has strong positive correlations in some tissues, including the kidney and brain. Furthermore, we discovered several strong negative correlations between hypomethylated CpG sites in T2 and genes expressed in brain, liver, and testis tissues, such as ZSCAN12P1_cg17849569, CYP1A1_cg12101586, CYP1A1_cg13570656, CYP1A1_cg22549041, and USP6_cg05918002. Finally, several strong positive correlations were found between hypomethylated CpG sites in T3 and genes expressed across various tissues. For example, AC009336.19_cg01152019 and HOXD4_cg01152019 exhibited strong positive correlations in the brain. However, significant negative correlations were also found in the brain and the kidney tissues, e.g., PPCS_cg06520846 and MOGAT1_cg22891868.

**Figure 2.**
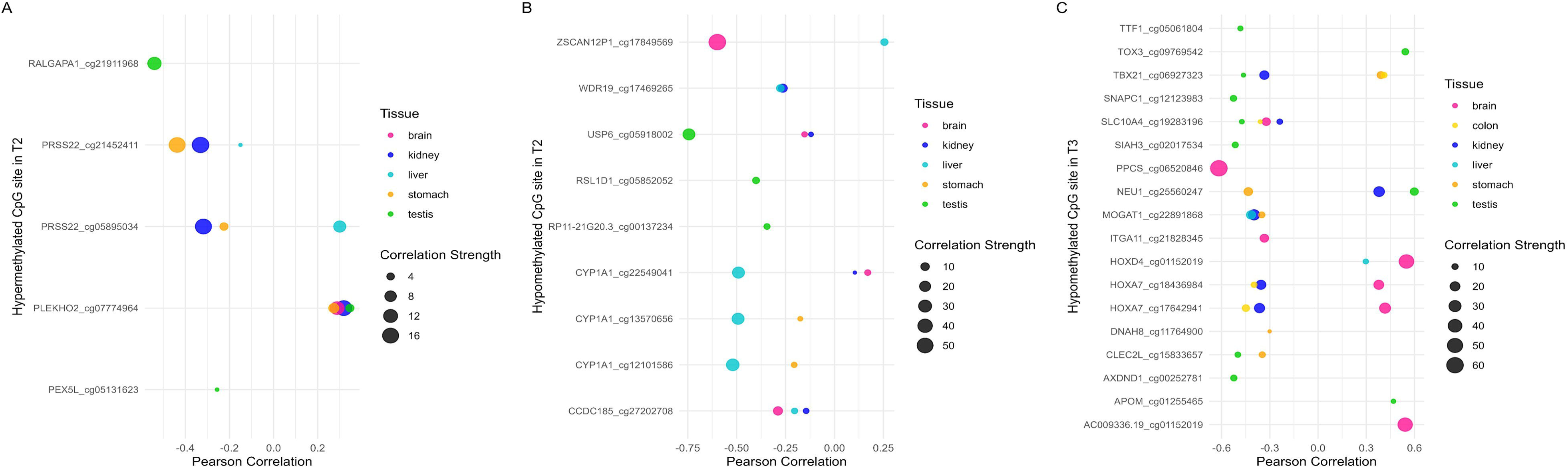
Dot plots showing potential regulatory associations between differentially methylated CpG sites and gene expression across six tissues (brain, colon, kidney, liver, stomach, and testis). (A) Hypermethylated CpG sites identified in T2, (B) Hypomethylated CpG sites identified in T2, and (C) Hypomethylated CpG sites identified in T3. Each dot represents the correlation strength of a CpG–gene pair in a specific tissue. The x-axis shows the Pearson correlation coefficient, dot color indicates tissue type, and dot size reflects the strength of association, represented as – log₁₀(p-value).

### Discovery of expression-quantitative trait methylation genes

Using the BIOS QTL browser data, we found an association between our DMPs and the expression of nearby genes. Our analysis identified 57 and 50 SNPs that were associated with hypermethylated and hypomethylated CpG sites in T2, respectively. The expression quantitative trait methylation gene (eQTM gene) identified from hypermethylated sites in T2 was *PDCD1*, whereas *USP6* and *ZNF232* were detected from hypomethylated sites. Furthermore, several SNPs were associated with hypermethylated (SNPs = 9) and hypomethylated (SNPs = 156) CpG sites identified in T3. Two eQTM genes (*MROH6* and *NAPRT1*) were detected from hypermethylated sites. On the other hand, hypomethylated sites in T3 were associated with several eQTM genes, including *RPS6KA2, MRPL10, HOXA1, RP4-728D4.2,* and *TFAP2E*.

Moreover, we linked the meQTL SNPs with known biological traits using the GWAS catalog. From hypomethylated sites in T2, the following traits were identified: body mass index (SNP: rs2470893), coffee consumption (rs2470893, rs2472304), caffeine metabolite measurement (rs2470893, rs35107470), and the phospholipid-to-total lipid ratio in very large HDLs (rs6465679). Only one SNP (rs10901802) associated with hypermethylated sites in T2 was related to high-density lipoprotein cholesterol measurement. On the other hand, several traits were found to be linked to hypomethylated sites at T3: BMI-adjusted waist circumference (rs68049170), diastolic blood pressure (rs62454420), body mass index (rs7274963), serine/threonine-protein kinase PAK 6 levels (rs2290768), apolipoprotein A1 levels (rs75988962), serum IgG glycosylation measurements (rs2093746), and high-sensitivity cardiac troponin I concentrations (rs8038368). We analyzed the clinical data collected from the QBiC to investigate coffee consumption traits. As a result, 31 out of 57 responders with GDM and 30 out of 63 non-GDM responders consumed instant coffee. While 30 of 57 GDM participants and 25 of 61 non-GDM participants consumed Arabic coffee with varying frequencies. However, the chi-square tests for coffee consumption revealed no significant differences between cases and controls, with p-values of 0.58 for instant coffee and 0.28 for Arabic coffee, respectively.

### Transcriptomic validation of the potential epigenetically dysregulated genes in GDM

Using publicly available gene expression profiling data (GSE255075), we validated several genes potentially dysregulated through DNA methylation changes. These genes included *USP6*, *RSL1D1*, *HOXD4*, *NEU1*, and *MROH6* as shown in Table 7. The transcriptomic data showed downregulation of the *USP6* gene in the placental tissue of the GDM women, while it was negatively correlated with the hypomethylated CpG site (cg05918002). Similarly, the *HOXD4* gene was downregulated; however, it was positively correlated with cg01152019. On the other hand, *RSL1D1*, *NEU1*, and *MROH6* were discovered to be upregulated in the placental tissues obtained from GDM pregnant women, with *MROH6* showing the highest significance (adjusted P-value of 8.68E-08) and the most substantial upregulation among all validated genes. Finally, the downregulation of *CARD8* was shown in the placental tissues, while we discovered the hypermethylation of the cg06392883 site, which is located upstream of the TSS region.

**Table 7:**
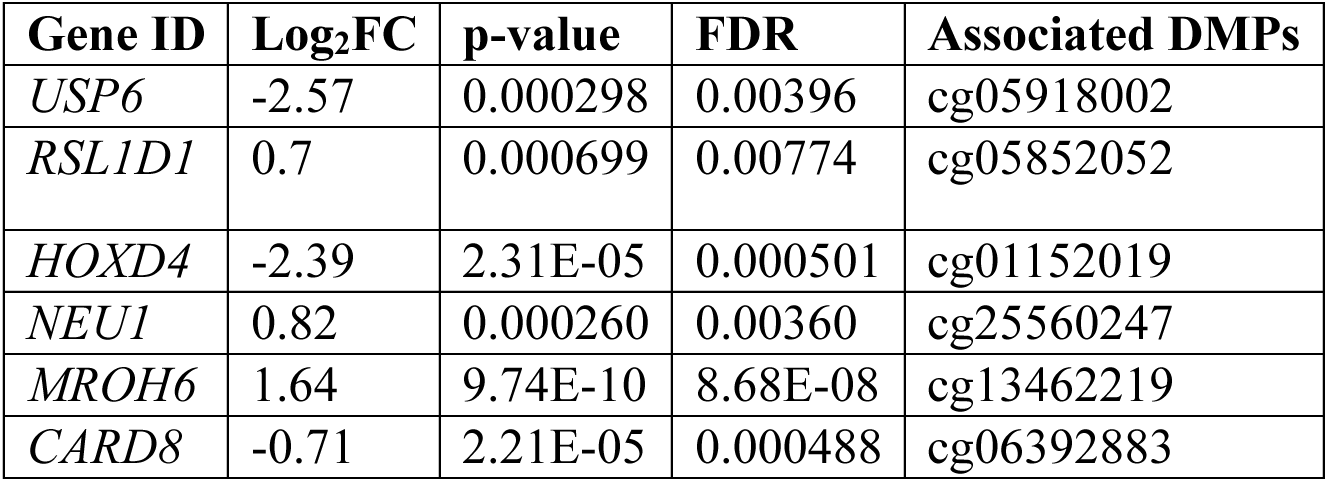
Expression-based validation of epigenetically identified genes in placental tissue from pregnant women with GDM. Abbreviations: FC, fold change; FDR, false discovery rate.

## Discussion

Gestational diabetes mellitus (GDM) is a common metabolic complication with first diagnosis between 24–28 weeks of pregnancy [39]. GDM is a complex disease with genetic, epigenetic, and environmental factors contributing to its etiology [5]. Previous studies have linked epigenetic modifications, particularly DNA methylation, to GDM [40, 41]. However, the relationship between DNA methylation changes and postnatal complications such as T2D and cardiovascular disorders in mothers remains underexplored, and the epigenetic mechanisms underlying GDM are not yet fully understood. The GDM prevalence is high, affecting 14% of pregnant women worldwide [39]. GDM prevalence in Qatar is even higher, as it reaches 31.6% due to the high incidence of obesity and diabetes in the population [42]. Thus, we conducted a genome-wide DNA methylation analysis in Qatar across the three trimesters of preganancy .

From this study, a significant difference in metabolic parameters between GDM cases and controls was identified, which aligns with the well-established associations between metabolic dysfunction and GDM. We identified differentially methylated sites and regions in the discovery cohort and validated our findings in the validation cohort. These CpG sites might influence GDM development. The downstream analysis of the validated CpG sites in T3 revealed pathways related to postnatal complications such as cardiomyopathy and diabetes mellitus. The hypomethylated sites in T3 also showed significant enrichment in blood, blood vessels, and liver. This observation may be partly because DNA methylation was measured in blood samples collected from pregnant women. The trait enrichment analysis using the EWAS toolkit highlighted the reflection of environmental and behavioral factors in DMPs. Furthermore, several meQTL SNPs had GWAS associations with BMI, phospholipid-to-total lipid ratio in very large HDL, diastolic blood pressure, high-sensitivity cardiac troponin I concentration, and other traits.

DNA methylation changes have been linked to GDM, primarily through studies focusing on offspring exposed to GDM *in utero*. Recent studies have explored the impact of GDM on DNA methylation signatures of pregnant women at two different time points. These studies performed an EWAS on a small sample of only 32 pregnant women, from whom peripheral blood samples were collected after GDM diagnosis at two distinct time points [29, 30]. Our study examined maternal DNA methylation patterns associated with GDM in a larger cohort, specifically in a Middle Eastern population where this condition is highly prevalent. In contrast to previous studies conducted on European and South Asian populations [29, 30], our analysis focuses on a Middle Eastern cohort and DNA methylation profiling was conducted across all three trimesters of pregnancy. Furthermore, we compared GDM associated DMPs to those identified in female patients with T2D collected at the Qatar BioBank. Here, we identified and validated several hypermethylated and hypomethylated CpG sites in T2 and T3. Unlike previous studies, our research highlighted pathways and CpG-associated traits related to cardiovascular disorders. Furthermore, we identified DMPs related to Type I diabetes, consistent with previous studies conducted in GDM mothers [29] and their children [43]. Our study revealed that several CpG sites may regulate the expression of genes associated with GDM, as indicated in Table 7. One of these genes is the *USP6* gene, which has been previously reported among the top upregulated differentially expressed genes in pregnant women with GDM and within the list of hub genes [44]. The *NEU1* gene was also discovered and validated using the GSE255075 dataset. This gene encodes neuraminidase-1, a membrane-associated sialidase that regulates insulin signaling by regulating the activity of insulin-induced insulin receptor tyrosine kinase. Previous studies have shown that human fibroblast cells lacking Neu1 will induce Neu3 sialidase activity when treated with Olanzapine, an antipsychotic agent that has been linked to insulin resistance. Furthermore, these studies indicate that the downregulation of Neu1 results in the impairment of insulin signaling, which contributes to insulin-resistant diabetes [45, 46]. We validated the dysregulation of a few other genes, including *RSL1D1*, *HOXD4*, and *MROH6*. These new genes have no previous relationship to GDM, and their role in GDM pathogenesis is yet to be discovered. Cytochrome P450 1A1 (CYP1A1) is a canonical target of the aryl hydrocarbon receptor (AhR), which is involved in the response to external stimuli such as environmental factors. We identified three hypomethylated CpG sites in T2 (cg12101586, cg13570656, and cg22549041) that might regulate the expression of this gene, *CYP1A1*, through promoter hypomethylation, as shown in Supplementary Table 9. Recent studies have shown that the upregulation of *CYP1A1* in male pancreatic islets triggered by stress responses associated with a high-fat diet might lead to insulin resistance and the development of β-cell dysfunction [47]. These findings suggest a potential role for AhR-CYP1A1 signaling in β-cell stress and glucose dysregulation [47]. Our analysis also revealed an association between the cg13570656 and rs2470893 SNPs, which were previously linked through GWAS to the *CYP1A1* gene and body mass index trait. Furthermore, during validation using the gene expression dataset GSE255075, *CYP11A1*, a member of the same gene family, was identified among the most highly dysregulated genes, with a log_2_ fold change (logFC) of 2.4 and an FDR-adjusted p-value of 1.30 × 10^-^¹⁴.

In this study, we also discovered common DNA methylation signatures that might contribute to the pathogenesis of GDM and T2D, and a few genes were identified, including *RGS4* and *CARD8*. The role of *RGS4* in regulating insulin release in T2D has been previously described, as it acts as a negative regulator of M3R (M3 muscarinic receptor) signaling in pancreatic β-cells. When M3R is activated, it stimulates Gq proteins to enhance glucose-stimulated insulin secretion via calcium signaling. RGS4 negatively regulates this pathway by accelerating the deactivation of Gq, thereby reducing M3R-mediated insulin release [48, 49]. *CARD8* has no direct impact on insulin release, but genetic variants in this gene have been previously reported to decrease the risk of diabetic nephropathy in T2D patients [50]. However, RGS4 and CARD8 have not been previously described in relation to GDM, and their role in this disease remains to be elucidated.

Furthermore, we discovered a few eQTM genes related to GDM and T2D via previously published cis-meQTLs [51]. These eQTM genes included *PDCD1, USP6*, *ZNF232, MROH6, NAPRT1*, *RPS6KA2, MRPL10, HOXA1, RP4-728D4.2,* and *TFAP2E*. The downregulation of *PDCD1* (PD-1) during the third trimester of pregnancy has been previously used as a marker of GDM incidence [52]. The associations of *RPS6KA2 and NAPRT1* with T2D were discovered in previous studies [53, 54]. The relationships of *ZNF232, MROH6, MRPL10*, *HOXA1*, *RP4-728D4.2*, and *TFAP2E* with GDM or diabetes have yet to be discovered.

Finally, our study showed how lifestyle and environmental factors can affect DNA methylation signatures during pregnancy. Few known biological traits identified in this study were related to lifestyle factors such as juice consumption, smoking, consuming nutritional supplements, caffeine consumption, BMI, and exercise. Other traits were related to environmental factors, including glucocorticoid exposure and *in-utero* arsenic exposure. Previous studies indicated the contribution of genetic and environmental factors to placental DNA methylation signatures [55] and how environmental stress during pregnancy also influences embryogenesis through DNA methylation changes [56]. Furthermore, smoking has been previously linked to DNA methylation changes in 15 CpG sites (including cg12101586 and cg22549041) associated with seven genes, *AHRR, MYO1G, GFI1, CYP1A1, CNTNAP2, KLF13*, and *ATP9A* [57]. In our study, two of these CpG sites were found to be hypomethylated in T2 and have a known regulatory effect on the same gene, *CYP1A1*. In a previous study, this gene was significantly upregulated in the placental tissues of smoking pregnant mothers compared to non-smoking pregnant women [58].

We acknowledge a few limitations in our current study. First, the number of samples collected during the first trimester of pregnancy was limited, and no T1 samples were obtained in the validation cohort. Second, analyzing genome-wide DNA methylation data without accompanying gene expression data limits validation and functional interpretation of the findings. Future longitudinal cohort studies that follow GDM women not just during pregnancy but also after delivery would be very important to assess epigenetic alterations and how they correlate with GDM progression and metabolic complications after delivery.

Overall, we identified differentially methylated CpG sites from genome-wide DNA methylation data when comparing GDM women to controls highlighting the potential role of DNA methylation changes in GDM pathogenesis. Our study suggests that DNA methylation signatures in pregnant women might reflect the influence of lifestyle and environmental factors before and during pregnancy. Our findings indicate that methylation status changes at specific sites might regulate the gene expression of several genes implicated in GDM and T2D. We provide new insights into the shared molecular mechanisms underlying these metabolic dysfunctions and highlight the importance of lifestyle and environmental factors on DNA methylation signatures. Additionally, we identified novel genes (*RSL1D1*, *HOXD4*, and *MROH6*) that may be epigenetically regulated in the context of GDM, expanding the current understanding of its genetic and epigenetic landscape. These results pave the way for future investigations into the functional consequences of epigenetic alterations affecting these genes and their potential utility in early prediction, risk assessment, and new treatment interventions for GDM.

## Contributions

LS: Writing – original draft, Writing – review & editing, and conducting experiments and data analysis. YB: Writing – original draft, Writing – review & editing. NEH: Supervision, Conceptualization, Funding, Writing – original draft, Writing – review & editing. JK, MD, and AS: Providing samples – conducting experiments. AQ: Writing – review & editing. NY: Writing – review & editing, Partial data analysis. EF: Providing samples, writing, review & editing. MB: Funding, Providing samples, Writing – review & editing. TF and NAC: Providing samples, Writing, review & editing. OC: Writing – review & editing. All authors have thoroughly reviewed and consented to the publication of the manuscript.

## Data Availability

The datasets generated and analyzed in this study are available from the corresponding author upon reasonable request, subject to institutional and ethical approvals.

## Acknowledgments

The authors would like to thank the QBiC participants, the research team, and all the Qatar Biobank personnel for contributing to this study.

