## Supplementary Table for "Maternal DNA Methylation Signatures of Gestational Diabetes across all Stages of Pregnancy"

**Supplementary Materials**

**Results**

**Table S1** GO enrichment analysis for validated hypermethylated CpG sites in T2

| Hypermethylated CpG sites | | | | | | |
| --- | --- | --- | --- | --- | --- | --- |
| ENTRY | ONTOLOGY | TERM | N | DE | P.DE | FDR |
| GO:0035269 | BP | protein O-linked mannosylation | 19 | 1 | 0.007562 | 1 |
| GO:0035268 | BP | protein mannosylation | 25 | 1 | 0.0094 | 1 |
| GO:0046716 | BP | muscle cell cellular homeostasis | 25 | 1 | 0.009542 | 1 |
| GO:0097502 | BP | mannosylation | 26 | 1 | 0.009709 | 1 |
| GO:0060045 | BP | positive regulation of cardiac muscle cell proliferation | 31 | 1 | 0.011588 | 1 |
| GO:0001580 | BP | detection of chemical stimulus involved in sensory perception of bitter taste | 36 | 1 | 0.01192 | 1 |
| GO:0055023 | BP | positive regulation of cardiac muscle tissue growth | 37 | 1 | 0.013297 | 1 |
| GO:0090162 | BP | establishment of epithelial cell polarity | 34 | 1 | 0.013301 | 1 |
| GO:0003382 | BP | epithelial cell morphogenesis | 34 | 1 | 0.01352 | 1 |
| GO:0050913 | BP | sensory perception of bitter taste | 41 | 1 | 0.013693 | 1 |
| GO:0050912 | BP | detection of chemical stimulus involved in sensory perception of taste | 42 | 1 | 0.014079 | 1 |
| GO:0060421 | BP | positive regulation of heart growth | 41 | 1 | 0.014879 | 1 |
| GO:0060043 | BP | regulation of cardiac muscle cell proliferation | 48 | 1 | 0.017706 | 1 |
| GO:0046622 | BP | positive regulation of organ growth | 53 | 1 | 0.019631 | 1 |
| GO:0050909 | BP | sensory perception of taste | 63 | 1 | 0.021988 | 1 |
| GO:0060038 | BP | cardiac muscle cell proliferation | 60 | 1 | 0.02263 | 1 |
| GO:0055021 | BP | regulation of cardiac muscle tissue growth | 67 | 1 | 0.024204 | 1 |
| GO:0060420 | BP | regulation of heart growth | 73 | 1 | 0.026495 | 1 |
| GO:0014855 | BP | striated muscle cell proliferation | 84 | 1 | 0.031861 | 1 |
| GO:0055017 | BP | cardiac muscle tissue growth | 91 | 1 | 0.033508 | 1 |
| GO:0060419 | BP | heart growth | 99 | 1 | 0.036523 | 1 |
| GO:0046620 | BP | regulation of organ growth | 102 | 1 | 0.03746 | 1 |
| GO:0006493 | BP | protein O-linked glycosylation | 100 | 1 | 0.037891 | 1 |

Abbreviations: BP, Biological Processes.

**Table S2** GO enrichment analysis for validated hypomethylated CpG sites in T2

| Hypomethylated CpG sites | | | | | | |
| --- | --- | --- | --- | --- | --- | --- |
| ENTRY | ONTOLOGY | TERM | N | DE | P.DE | FDR |
| GO:1903772 | BP | regulation of viral budding via host ESCRT complex | 3 | 1 | 0.001318 | 1 |
| GO:1903774 | BP | positive regulation of viral budding via host ESCRT complex | 3 | 1 | 0.001318 | 1 |
| GO:0043328 | BP | protein transport to vacuole involved in ubiquitin-dependent protein catabolic process via the multivesicular body sorting pathway | 15 | 1 | 0.004209 | 1 |
| GO:0000470 | BP | maturation of LSU-rRNA | 28 | 1 | 0.005529 | 1 |
| GO:0032511 | BP | late endosome to vacuole transport via multivesicular body sorting pathway | 26 | 1 | 0.005751 | 1 |
| GO:0039702 | BP | viral budding via host ESCRT complex | 21 | 1 | 0.005842 | 1 |
| GO:1903902 | BP | positive regulation of viral life cycle | 24 | 1 | 0.007016 | 1 |
| GO:0046755 | BP | viral budding | 24 | 1 | 0.007089 | 1 |
| GO:0045324 | BP | late endosome to vacuole transport | 36 | 1 | 0.008319 | 1 |
| GO:0036258 | BP | multivesicular body assembly | 31 | 1 | 0.008433 | 1 |
| GO:0019076 | BP | viral release from host cell | 28 | 1 | 0.008447 | 1 |
| GO:0035891 | BP | exit from host cell | 28 | 1 | 0.008447 | 1 |
| GO:0036257 | BP | multivesicular body organization | 32 | 1 | 0.008885 | 1 |
| GO:0043162 | BP | ubiquitin-dependent protein catabolic process via the multivesicular body sorting pathway | 36 | 1 | 0.009695 | 1 |
| GO:0019068 | BP | virion assembly | 33 | 1 | 0.010035 | 1 |
| GO:0032509 | BP | endosome transport via multivesicular body sorting pathway | 40 | 1 | 0.010231 | 1 |
| GO:0072657 | BP | protein localization to membrane | 605 | 2 | 0.011258 | 1 |
| GO:0090148 | BP | membrane fission | 44 | 1 | 0.012509 | 1 |
| GO:2000772 | BP | regulation of cellular senescence | 55 | 1 | 0.012929 | 1 |
| GO:0071985 | BP | multivesicular body sorting pathway | 47 | 1 | 0.013043 | 1 |
| GO:0006623 | BP | protein targeting to vacuole | 48 | 1 | 0.013958 | 1 |
| GO:0051668 | BP | localization within membrane | 695 | 2 | 0.01503 | 1 |
| GO:0042273 | BP | ribosomal large subunit biogenesis | 73 | 1 | 0.015787 | 1 |
| GO:0048524 | BP | positive regulation of viral process | 64 | 1 | 0.017767 | 1 |
| GO:0006940 | BP | regulation of smooth muscle contraction | 60 | 1 | 0.018778 | 1 |
| GO:0072666 | BP | establishment of protein localization to vacuole | 65 | 1 | 0.019125 | 1 |
| GO:0008104 | BP | protein localization | 2480 | 3 | 0.02465 | 1 |
| GO:0070727 | BP | cellular macromolecule localization | 2491 | 3 | 0.024968 | 1 |
| GO:0007032 | BP | endosome organization | 96 | 1 | 0.025051 | 1 |
| GO:0072665 | BP | protein localization to vacuole | 88 | 1 | 0.026227 | 1 |
| GO:0006612 | BP | protein targeting to membrane | 126 | 1 | 0.030554 | 1 |
| GO:0090398 | BP | cellular senescence | 109 | 1 | 0.031186 | 1 |
| GO:0006939 | BP | smooth muscle contraction | 108 | 1 | 0.034953 | 1 |
| GO:1903900 | BP | regulation of viral life cycle | 142 | 1 | 0.035558 | 1 |
| GO:0033036 | BP | macromolecule localization | 2974 | 3 | 0.03877 | 1 |
| GO:0050792 | BP | regulation of viral process | 167 | 1 | 0.042007 | 1 |
| GO:0006937 | BP | regulation of muscle contraction | 167 | 1 | 0.048324 | 1 |
| GO:0044085 | BP | cellular component biogenesis | 3303 | 3 | 0.048814 | 1 |
| GO:0007034 | BP | vacuolar transport | 170 | 1 | 0.049281 | 1 |

Abbreviations: BP, Biological Processes.

**Table S3** GO enrichment analysis for validated hypermethylated CpG sites in T3

| Hypermethylated CpG sites | | | | | |
| --- | --- | --- | --- | --- | --- |
| ENTRY | ONTOLOGY | TERM | N | DE | P.DE |
| GO:0005975 | BP | carbohydrate metabolic process | 556 | 1 | 0.035001 |

Abbreviations: BP, Biological Processes.

**Table S4** GO enrichment analysis for validated hypomethylated CpG sites in T3

| Hypomethylated CpG sites | | | | | | |
| --- | --- | --- | --- | --- | --- | --- |
|  | ONTOLOGY | TERM | N | DE | P.DE | FDR |
| GO:0098874 | BP | spike train | 1 | 1 | 0.001617 | 1 |
| GO:0099608 | BP | regulation of action potential firing pattern | 1 | 1 | 0.001617 | 1 |
| GO:0071348 | BP | cellular response to interleukin-11 | 1 | 1 | 0.00172 | 1 |
| GO:0006540 | BP | glutamate decarboxylation to succinate | 2 | 1 | 0.002241 | 1 |
| GO:2000329 | BP | negative regulation of T-helper 17 cell lineage commitment | 2 | 1 | 0.002801 | 1 |
| GO:2000552 | BP | negative regulation of T-helper 2 cell cytokine production | 4 | 1 | 0.002951 | 1 |
| GO:0071105 | BP | response to interleukin-11 | 2 | 1 | 0.003375 | 1 |
| GO:1903215 | BP | negative regulation of protein targeting to mitochondrion | 5 | 1 | 0.005379 | 1 |
| GO:0002296 | BP | T-helper 1 cell lineage commitment | 4 | 1 | 0.005804 | 1 |
| GO:0070072 | BP | vacuolar proton-transporting V-type ATPase complex assembly | 6 | 1 | 0.006065 | 1 |
| GO:0006105 | BP | succinate metabolic process | 7 | 1 | 0.006632 | 1 |
| GO:1903748 | BP | negative regulation of establishment of protein localization to mitochondrion | 7 | 1 | 0.007477 | 1 |
| GO:0009449 | BP | gamma-aminobutyric acid biosynthetic process | 6 | 1 | 0.007537 | 1 |
| GO:0072675 | BP | osteoclast fusion | 6 | 1 | 0.008003 | 1 |
| GO:0035744 | BP | T-helper 1 cell cytokine production | 7 | 1 | 0.008262 | 1 |
| GO:2000554 | BP | regulation of T-helper 1 cell cytokine production | 7 | 1 | 0.008262 | 1 |
| GO:2000556 | BP | positive regulation of T-helper 1 cell cytokine production | 7 | 1 | 0.008262 | 1 |
| GO:0006538 | BP | glutamate catabolic process | 8 | 1 | 0.008528 | 1 |
| GO:0070070 | BP | proton-transporting V-type ATPase complex assembly | 8 | 1 | 0.008976 | 1 |
| GO:0042796 | BP | snRNA transcription by RNA polymerase III | 9 | 1 | 0.009491 | 1 |
| GO:2000328 | BP | regulation of T-helper 17 cell lineage commitment | 8 | 1 | 0.009671 | 1 |
| GO:0009448 | BP | gamma-aminobutyric acid metabolic process | 8 | 1 | 0.009863 | 1 |
| GO:0002725 | BP | negative regulation of T cell cytokine production | 10 | 1 | 0.009931 | 1 |
| GO:0072674 | BP | multinuclear osteoclast differentiation | 8 | 1 | 0.010479 | 1 |
| GO:0006651 | BP | diacylglycerol biosynthetic process | 9 | 1 | 0.010522 | 1 |
| GO:0048304 | BP | positive regulation of isotype switching to IgG isotypes | 10 | 1 | 0.010759 | 1 |
| GO:2000320 | BP | negative regulation of T-helper 17 cell differentiation | 10 | 1 | 0.012627 | 1 |
| GO:0035745 | BP | T-helper 2 cell cytokine production | 12 | 1 | 0.013051 | 1 |
| GO:2000551 | BP | regulation of T-helper 2 cell cytokine production | 12 | 1 | 0.013051 | 1 |
| GO:1990034 | BP | calcium ion export across plasma membrane | 11 | 1 | 0.013183 | 1 |
| GO:0010454 | BP | negative regulation of cell fate commitment | 10 | 1 | 0.013569 | 1 |
| GO:0036295 | BP | cellular response to increased oxygen levels | 12 | 1 | 0.013679 | 1 |
| GO:2000317 | BP | negative regulation of T-helper 17 type immune response | 11 | 1 | 0.013778 | 1 |
| GO:0099566 | BP | regulation of postsynaptic cytosolic calcium ion concentration | 10 | 1 | 0.014301 | 1 |
| GO:0048302 | BP | regulation of isotype switching to IgG isotypes | 14 | 1 | 0.015539 | 1 |
| GO:0002829 | BP | negative regulation of type 2 immune response | 15 | 1 | 0.016043 | 1 |
| GO:0048291 | BP | isotype switching to IgG isotypes | 15 | 1 | 0.016673 | 1 |
| GO:0042795 | BP | snRNA transcription by RNA polymerase II | 16 | 1 | 0.016857 | 1 |
| GO:0002827 | BP | positive regulation of T-helper 1 type immune response | 17 | 1 | 0.017297 | 1 |
| GO:0070071 | BP | proton-transporting two-sector ATPase complex assembly | 16 | 1 | 0.017867 | 1 |
| GO:0043649 | BP | dicarboxylic acid catabolic process | 17 | 1 | 0.018237 | 1 |
| GO:0061430 | BP | bone trabecula morphogenesis | 13 | 1 | 0.018284 | 1 |
| GO:0000413 | BP | protein peptidyl-prolyl isomerization | 17 | 1 | 0.018484 | 1 |
| GO:0035743 | BP | CD4-positive, alpha-beta T cell cytokine production | 18 | 1 | 0.019853 | 1 |
| GO:0038065 | BP | collagen-activated signaling pathway | 15 | 1 | 0.020146 | 1 |
| GO:0051014 | BP | actin filament severing | 16 | 1 | 0.020268 | 1 |
| GO:0072540 | BP | T-helper 17 cell lineage commitment | 17 | 1 | 0.020316 | 1 |
| GO:0170043 | BP | non-proteinogenic amino acid biosynthetic process | 17 | 1 | 0.020561 | 1 |
| GO:0048172 | BP | regulation of short-term neuronal synaptic plasticity | 16 | 1 | 0.020666 | 1 |
| GO:0002281 | BP | macrophage activation involved in immune response | 19 | 1 | 0.020795 | 1 |
| GO:0045623 | BP | negative regulation of T-helper cell differentiation | 17 | 1 | 0.021242 | 1 |
| GO:0009301 | BP | snRNA transcription | 20 | 1 | 0.021659 | 1 |
| GO:0006071 | BP | glycerol metabolic process | 20 | 1 | 0.022478 | 1 |
| GO:1905146 | BP | lysosomal protein catabolic process | 20 | 1 | 0.02452 | 1 |
| GO:0019400 | BP | alditol metabolic process | 23 | 1 | 0.025962 | 1 |
| GO:0002295 | BP | T-helper cell lineage commitment | 21 | 1 | 0.025983 | 1 |
| GO:2000319 | BP | regulation of T-helper 17 cell differentiation | 22 | 1 | 0.026392 | 1 |
| GO:0009065 | BP | glutamine family amino acid catabolic process | 24 | 1 | 0.027277 | 1 |
| GO:0043371 | BP | negative regulation of CD4-positive, alpha-beta T cell differentiation | 21 | 1 | 0.027436 | 1 |
| GO:0045063 | BP | T-helper 1 cell differentiation | 23 | 1 | 0.028182 | 1 |
| GO:0002710 | BP | negative regulation of T cell mediated immunity | 27 | 1 | 0.028295 | 1 |
| GO:0007042 | BP | lysosomal lumen acidification | 24 | 1 | 0.028363 | 1 |
| GO:0015721 | BP | bile acid and bile salt transport | 29 | 1 | 0.028803 | 1 |
| GO:0043373 | BP | CD4-positive, alpha-beta T cell lineage commitment | 24 | 1 | 0.029329 | 1 |
| GO:0007039 | BP | protein catabolic process in the vacuole | 24 | 1 | 0.02954 | 1 |
| GO:0002726 | BP | positive regulation of T cell cytokine production | 25 | 1 | 0.029861 | 1 |
| GO:0032703 | BP | negative regulation of interleukin-2 production | 27 | 1 | 0.029991 | 1 |
| GO:0002825 | BP | regulation of T-helper 1 type immune response | 30 | 1 | 0.030546 | 1 |
| GO:0036296 | BP | response to increased oxygen levels | 25 | 1 | 0.030772 | 1 |
| GO:0051016 | BP | barbed-end actin filament capping | 25 | 1 | 0.03181 | 1 |
| GO:0006929 | BP | substrate-dependent cell migration | 26 | 1 | 0.031847 | 1 |
| GO:0002363 | BP | alpha-beta T cell lineage commitment | 26 | 1 | 0.032087 | 1 |
| GO:0045830 | BP | positive regulation of isotype switching | 28 | 1 | 0.032125 | 1 |
| GO:0046339 | BP | diacylglycerol metabolic process | 27 | 1 | 0.033864 | 1 |
| GO:0046639 | BP | negative regulation of alpha-beta T cell differentiation | 26 | 1 | 0.035123 | 1 |
| GO:0043369 | BP | CD4-positive or CD8-positive, alpha-beta T cell lineage commitment | 28 | 1 | 0.035389 | 1 |
| GO:0018208 | BP | peptidyl-proline modification | 31 | 1 | 0.035593 | 1 |
| GO:1903779 | BP | regulation of cardiac conduction | 28 | 1 | 0.035733 | 1 |
| GO:0006814 | BP | sodium ion transport | 242 | 2 | 0.036145 | 1 |
| GO:0006536 | BP | glutamate metabolic process | 32 | 1 | 0.036289 | 1 |
| GO:0002828 | BP | regulation of type 2 immune response | 33 | 1 | 0.036721 | 1 |
| GO:0035751 | BP | regulation of lysosomal lumen pH | 32 | 1 | 0.037193 | 1 |
| GO:2000515 | BP | negative regulation of CD4-positive, alpha-beta T cell activation | 30 | 1 | 0.037405 | 1 |
| GO:0045684 | BP | positive regulation of epidermis development | 32 | 1 | 0.037956 | 1 |
| GO:2000316 | BP | regulation of T-helper 17 type immune response | 32 | 1 | 0.038244 | 1 |
| GO:0002719 | BP | negative regulation of cytokine production involved in immune response | 34 | 1 | 0.039257 | 1 |
| GO:0072539 | BP | T-helper 17 cell differentiation | 33 | 1 | 0.039783 | 1 |
| GO:0002369 | BP | T cell cytokine production | 37 | 1 | 0.041078 | 1 |
| GO:0002724 | BP | regulation of T cell cytokine production | 37 | 1 | 0.041078 | 1 |
| GO:0042113 | BP | B cell activation | 275 | 2 | 0.04216 | 1 |
| GO:0007035 | BP | vacuolar acidification | 36 | 1 | 0.042449 | 1 |
| GO:0002360 | BP | T cell lineage commitment | 34 | 1 | 0.042457 | 1 |
| GO:0045191 | BP | regulation of isotype switching | 37 | 1 | 0.042559 | 1 |
| GO:0071354 | BP | cellular response to interleukin-6 | 40 | 1 | 0.044156 | 1 |
| GO:0002366 | BP | leukocyte activation involved in immune response | 299 | 2 | 0.045234 | 1 |
| GO:0002714 | BP | positive regulation of B cell mediated immunity | 41 | 1 | 0.046297 | 1 |
| GO:0002891 | BP | positive regulation of immunoglobulin mediated immune response | 41 | 1 | 0.046297 | 1 |
| GO:0002263 | BP | cell activation involved in immune response | 303 | 2 | 0.046616 | 1 |
| GO:0002183 | BP | cytoplasmic translational initiation | 42 | 1 | 0.046709 | 1 |
| GO:0042092 | BP | type 2 immune response | 41 | 1 | 0.047028 | 1 |
| GO:0002701 | BP | negative regulation of production of molecular mediator of immune response | 44 | 1 | 0.048794 | 1 |
| GO:0106056 | BP | regulation of calcineurin-mediated signaling | 41 | 1 | 0.0488 | 1 |
| GO:0070741 | BP | response to interleukin-6 | 44 | 1 | 0.04928 | 1 |
| GO:0051693 | BP | actin filament capping | 40 | 1 | 0.04982 | 1 |

Abbreviations: BP, Biological Processes.

**Table S5** EWAS toolkit traits associated with hypermethylated sites investigated in T2

| Trait | Odds Ratio | p-value | DMC | Background |
| --- | --- | --- | --- | --- |
| autoimmune diseases | 62.628 | 2.47E-16 | 1 | 372 |
| juice consumption | 33.727 | 2.20E-12 | 2 | 749 |
| atherosclerosis | 17.742 | 4.05E-12 | 2 | 2202 |
| leukoaraiosis (LA) | 40.969 | 4.79E-12 | 1 | 531 |
| down syndrome | 5.745 | 3.00E-09 | 6 | 14649 |
| ancestry | 5.255 | 2.19E-06 | 5 | 10618 |
| papillary thyroid carcinoma | 6.26 | 4.61E-06 | 3 | 6073 |
| smoking | 3.545 | 3.34E-05 | 6 | 20798 |
| preterm birth | 4.429 | 5.11E-05 | 4 | 10662 |
| psoriasis | 6.136 | 9.24E-05 | 3 | 4138 |
| vitamin B12 supplement | 16.744 | 1.27E-04 | 1 | 589 |
| glucocorticoid exposure | 5.961 | 2.87E-04 | 2 | 3468 |
| primary Sjögren's Syndrome (pSS) | 5.896 | 3.06E-04 | 2 | 3526 |
| Kabuki syndrome (KS) | 7.509 | 7.36E-04 | 2 | 1891 |
| cognitive function | 16.419 | 9.56E-04 | 1 | 391 |
| childhood stress | 13.196 | 1.78E-03 | 1 | 550 |
| oral squamous cell carcinoma (OSCC) | 3.068 | 1.88E-03 | 5 | 14095 |
| aging | 2.395 | 2.90E-03 | 7 | 31184 |
| exercise | 6.646 | 3.76E-03 | 1 | 1652 |

**Table S6** EWAS toolkit traits associated with hypomethylated sites investigated in T2

| Trait | Odds Ratio | p-value | DMC | Background |
| --- | --- | --- | --- | --- |
| ancestry | 14.566 | 2.67E-11 | 7 | 10618 |
| maternal smoking | 16.411 | 1.13E-07 | 4 | 6153 |
| leukoaraiosis (LA) | 91.59 | 1.88E-07 | 2 | 531 |
| colorectal laterally spreading tumor | 8.906 | 1.11E-04 | 3 | 8245 |
| estimated glomerular filtration rate (eGFR) | 97.648 | 2.24E-04 | 1 | 243 |
| Behcet's disease | 11.032 | 6.65E-04 | 2 | 4364 |
| In-utero arsenic exposure | 36.16 | 1.59E-03 | 1 | 655 |
| neurodevelopmental presentations and congenital anomalies (ND/CAs) | 27.656 | 2.68E-03 | 1 | 856 |
| prostate cancer | 6.791 | 3.80E-03 | 2 | 7047 |
| smoking | 3.372 | 4.98E-03 | 4 | 20798 |
| systemic lupus erythematosus (SLE) | 6.087 | 5.54E-03 | 2 | 7848 |

**Table S7** EWAS toolkit traits associated with hypomethylated sites investigated in T3

| Trait | Odds Ratio | p-value | DMC | Background |
| --- | --- | --- | --- | --- |
| aging | 5.157 | 1.84E-20 | 25 | 31184 |
| Nicolaides–Baraitser syndrome (NCBRS) | 33.007 | 1.67E-13 | 1 | 356 |
| juice consumption | 20.538 | 3.19E-12 | 1 | 749 |
| maternal smoking | 6.711 | 1.13E-11 | 6 | 6153 |
| SETD1B-related syndrome | 7.036 | 1.36E-07 | 5 | 2721 |
| Claes-Jensen syndrome | 10.382 | 1.92E-06 | 2 | 1050 |
| blood protein biomarker levels | 24.648 | 2.71E-06 | 1 | 168 |
| soluble tumor necrosis factor receptor 2 (sTNFR2) levels in plasma | 24.648 | 2.71E-06 | 1 | 168 |
| B Acute Lymphoblastic Leukemia with t(1;19)(q23;p13.3); E2A-PBX1 (TCF3-PBX1) | 2.95 | 2.82E-06 | 10 | 21099 |
| childhood stress | 13.587 | 8.10E-06 | 1 | 550 |
| fetal alcohol spectrum disorder (FASD) | 11.015 | 1.21E-04 | 1 | 571 |
| myalgic encephalomyelitis/chronic fatigue syndrome | 3.58 | 1.52E-04 | 4 | 6500 |
| nasopharyngeal carcinoma | 7.666 | 1.84E-04 | 1 | 1068 |
| Werner syndrome | 7.586 | 1.95E-04 | 1 | 1088 |
| smoking | 2.326 | 6.09E-04 | 9 | 20798 |
| Hyperdiploid B Acute Lymphoblastic Leukemia | 4.116 | 1.06E-03 | 2 | 2798 |
| body mass index (BMI) | 4.725 | 2.19E-03 | 1 | 1797 |
| colorectal laterally spreading tumor | 2.734 | 2.48E-03 | 4 | 8245 |
| autoimmune diseases | 8.708 | 5.48E-03 | 1 | 372 |
| respiratory allergies (RA) | 7.44 | 8.40E-03 | 1 | 485 |

**Figure Legends**

Figure S1. Venn diagram showing the validated CpG sites specifically in T2 and T3 in both the QBiC and WWRC cohorts.
