## Supplementary figures and images for "Maternal DNA Methylation Signatures of Gestational Diabetes across all Stages of Pregnancy"

### Supplementary Figure 1

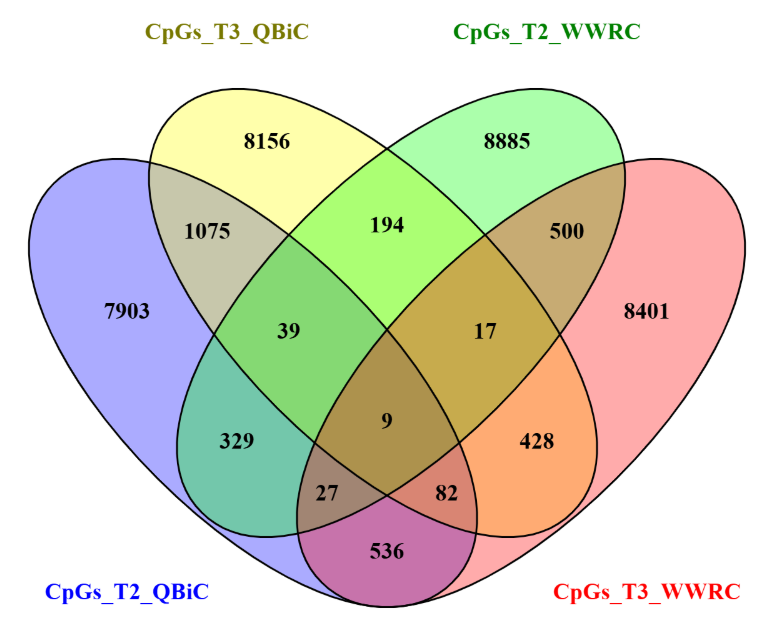
